# Pre-stimulus low-alpha frontal networks characterize pareidolias in Parkinson’s disease

**DOI:** 10.1101/2020.12.09.20246850

**Authors:** Gajanan S. Revankar, Yuta Kajiyama, Noriaki Hattori, Tetsuya Shimokawa, Tomohito Nakano, Masahito Mihara, Etsuro Mori, Hideki Mochizuki

## Abstract

**Background:** Parkinson’s disease (PD) patients susceptible to visual hallucinations experience perceptual deficits in the form of pareidolias. While pareidolias necessitate top-down modulation of visual processing, the cortical dynamics of internally generated perceptual priors on pareidolic misperceptions is unknown.

**Objectives:** To study pre-stimulus related EEG spectral and network abnormalities in PD patients experiencing pareidolias.

**Methods:** 21 PD in-patients and 10 age-matched healthy controls were evaluated. Neuropsychological assessments included tests for cognition, attention and executive functions. To evoke and quantify pareidolias, participants performed the noise pareidolia test (NPT) with simultaneous EEG recording. PD patients were sub-divided into two groups - those with high pareidolia counts (N=10) and those without (N=11). EEG was analyzed 1000ms before stimulus presentation in the spectral domain (theta, low-alpha and high-alpha frequencies) with corresponding graph networks that evaluated small-world properties, efficiency and centrality measures. Statistical analysis included ANCOVA and multiple regression to evaluate the differences.

**Results:** PD group with high pareidolias were older with lower scores on neuropsychological tests. Their pre-stimulus EEG low-alpha band showed a tendency towards higher frontal activity (p=0.06). Graph networks showed increased normalized clustering coefficient (p=0.05), higher local parietal cortex efficiency (p=0.049) and lower frontal degree centrality (p=0.005). These network indices correlated positively to patients’ pareidolia scores.

**Conclusion:** Pareidolias in PD are a consequence of an abnormal top-down modulation of visual processing which are defined by their frontal low-alpha spectral and network alterations in the pre-stimulus phase due to a dissonance between patients’ internally generated mental-processing with external stimuli.

## Introduction

Parkinson’s disease (PD) is predominantly a motor neurodegenerative disease, but those affected frequently show non-motor symptoms of psychosis such as mood disorders, impulsivity and hallucinations(1). Specifically, visual hallucinations (VH) have a strong prevalence in PD and contribute adversely to motor symptoms(2,3). Notably, VH are often preceded by minor hallucinations(4) wherein patients develop misperceptions of inanimate forms, objects or shapes(1). These visuo-perceptual deficits commonly occur in the form of ‘pareidolias’ and serve as precursors in identifying hallucinatory tendencies in PD patients(5). Several studies have previously shown a commonality of features between pareidolia and VH in patient populations(5–8) making it ideal to quantify pareidolias as a proxy for VH.

Clinically, pareidolias can be quantified using the noise pareidolia test (NPT), a neuropsychological test wherein ambiguous patterns are presented to evoke illusionary responses in PD or dementia with Lewy body patients(5,9). Pareidolias evoked on the NPT inherently prompt a visuo-perceptual demand(5) that requires reorienting attention depending on the salience revealed by the target. Recently, in a multimodal electroencephalography (EEG) experiment, we showed that PD patients exhibited abnormal top-down modulation of visual information *‘during’* pareidolic manifestations(10), suggestive of deficits in matching external sensory information with prior knowledge(11). However, considering the impact of abnormal priors in the causation of pareidolias(12,13), it is yet to be determined how neural information ‘*preceding’* stimulus onset influences perception in those susceptible to pareidolias. To that end, we investigated how pre-stimulus cortical activity affects visual processing in PD patients during the performance of the NPT.

Prior studies on pre-stimulus oscillations have detailed the causal role of alpha frequencies in shaping and detecting sub-threshold stimuli(14). The voluntary visual attentional stream, defined by the connections between the frontal and parietal cortices, reportedly have a strong impact of alpha band oscillations in discriminating ambiguous targets(15) as well as prediction of perceptual decisions(16). Within the domains of attention and working memory, cortical alpha has been variously described under different terminologies such as mind-wandering(17), creative ideation(18) or mental imagery(19), all to emphasize the influence of subjects’ internally-generated mental operations(20) in the absence of bottom-up sensory information. Considering that the communication between these exo- and endogenous attentional systems is impaired in PD patients with VH(21), low-frequency cortical oscillations may have a crucial role in the generation and maintenance of misperceptual tendencies. Specifically in PD without dementia, an increase in theta and low-alpha power is reported in resting-state recordings(22,23), with stimulus dependent studies reinforcing the importance of alpha bands(24). As symptoms and severity worsen, theta-pre-alpha rhythms in the frontal cortex has been shown to predict the cognitive decline(25) and is associated with hallucinations and/or delusions(26). Regardless, the visuo-perceptual dynamics in PD patients prior to stimulus onset is currently unknown.

Whereas EEG studies conventionally use event-related and/or time-frequency analysis to study stimulus related effects, application of graph theory concepts to EEG data provides an efficient modality to examine alterations in higher-level whole-network brain areas(27). Given that pareidolias directly relate to a failure of attentional(10) and perceptual decision-making(6), the involvement of multiple attentional network domains(28) suggestive of a global cortical dysfunction cannot be overlooked. Therefore, evaluating pre-stimulus graph network metrics is speculated to clarify the abnormal network organization in such patients. Prior studies on PD have demonstrated decreased ‘small world’ properties, network efficiency and centrality measures when compared to healthy controls, with the outcomes worsening further in PD with dementias(29–31). Accordingly, in our present study, we will focus on deriving frequency specific graph metrics associated with pareidolias and their impact on visuo-perception.

In summary, we hypothesized that PD patients with pareidolic tendencies demonstrate aberrant low-frequency oscillations with reciprocal network insufficiencies, addressed in an experimental paradigm where decisions are critically dependent on the individual’s prior expectation of the stimulus. To achieve these aims, we performed an exploratory analysis on pre-stimulus intervals in PD patients and healthy controls (HC) to study the global and local (fronto-parietal) network dynamics using EEG spectral and graph-theory indices when participants performed the NPT. The results of our current work are expected to reinforce the findings of our prior study(10) and further our understanding on the effects of pre-stimulus cortical activity that shape post-stimulus pareidolic misperceptions.

## Materials and Methods

### General information and recording procedures

We analyzed a dataset of 21 PD in-patients [age 70.4y±8.5 (Mean, SD)] and age-matched 12 healthy controls (69.4y±8.5) whose details are published previously(10). Notably, PD patients with dementia were excluded (MMSE scores <24) and testing was done during ‘ON’ state of PD medication. All participants had normal or corrected-to-normal vision. Neuropsychological assessments relevant to the study were conducted by a trained psychologist, independent of experimenters who performed EEG. These included the Japanese adult reading test (JART), frontal assessment battery (FAB), Montreal cognitive assessment (MoCA) and the short-form of Benton’s judgment of line orientation test (JLO, Form H). At the outset, we classified the patients based on the clinical noise pareidolia test (NPT), a standardized paper-based test to evoke and quantify pareidolias(32). The test comprises of black and white patches of visual noise with certain images containing faces. Correct responses refer to appropriate discrimination of faces from noise. However, when faces are identified in stimuli without a face, such responses are marked as pareidolias. Unidentified faces are categorized as missed responses. The test uses misperceptual cutoff scores of 0-1 as non-pareidolics and ≥ 2 as pareidolics. Of the 21 PD patients, 10 exhibited high pareidolia scores, henceforth classified as PD pareidolia type (PDP), and the rest were classified as PD non-pareidolic type (PDnP). There were 2 dropouts within the HC group (due to technical difficulties) bringing the total number of tested participants to HC=10, PDnP=11 and PDP=10 for final EEG analysis. A summary of the participant details is shown in Table-1. Informed consent was obtained from all subjects and in accordance with the ethical standards of the Declaration of Helsinki, the institutional review board cleared the protocol for the study (approval number 18136).

For the EEG experiment, participants were shown 80 NPT images on a computer monitor, 60 with visual noise and 20 with faces embedded in them (Supplementary Fig. 1). The faces on the NPT are low-information, 2-toned images which requires configural knowledge based facial-feature processing(33). Cutoffs were appropriately increased. Participants were seated upright at a distance of 80cms from the monitor display which had images subtending a size of 700×700 pixels (width x height). Participants verbally responded to each image within 30s of stimulus presentation and the experimenter recorded the responses since patients’ tremor severity may have prevented reliable manual responses. EEG data were recorded on a 32-channel Biosemi, ActiveTwo system at 2kHz sampling rate on ActiView software (LabVIEW). EEG recordings were re-referenced to average of all the electrode channels, pass filtered between 1Hz and 45Hz and down-sampled to 500Hz for independent component analysis. Final EEG data was then reconstructed after independent components responsible for heartbeat, eye-blinks, and muscle related artifacts were removed using visual inspection.

**Figure 1.**
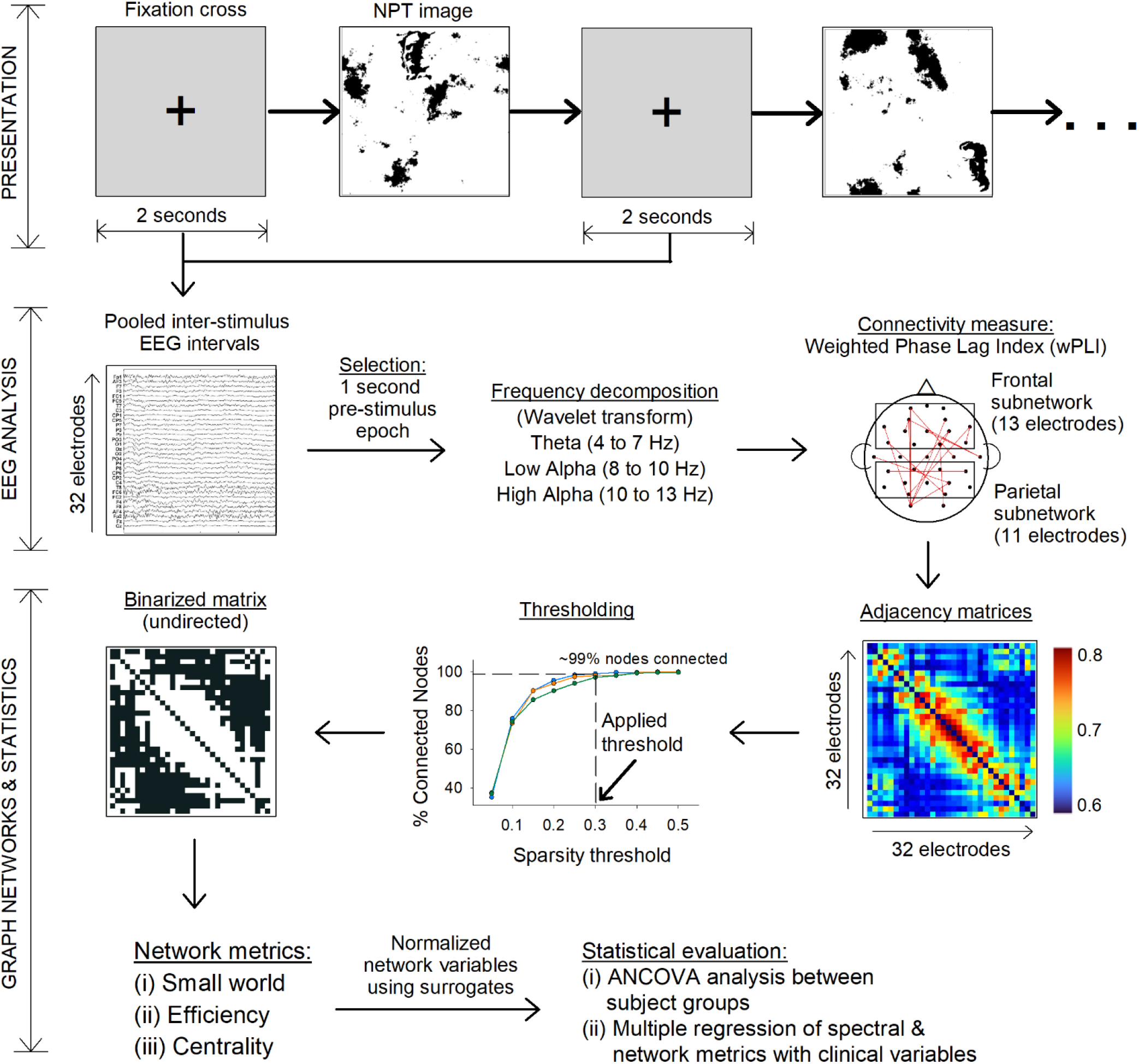
Overview of presentation, recording and analysis methods. Legend Fig.1: NPT image = noise pareidolia test image, EEG = electroencephalography, ANCOVA = analysis of covariance. The workflow was divided into three parts as shown on left margins: Presentation of stimuli, EEG analysis and Graph network analysis. Presentation of stimuli comprised of 80 NPT images, each preceded by a fixation cross for 2 seconds which served as epochs for EEG analysis. EEG analysis involved preprocessing and wPLI calculation for theta, low-alpha and high-alpha band frequencies. Finally graph network were calculated based on binarized matrices obtained using wPLI as edges and EEG electrodes as nodes. Line-plot shown in thresholding represent the 3 groups that were tested. Statistical comparisons were subsequently performed for spectral and network measures between the three participant groups.

**Figure 2.**
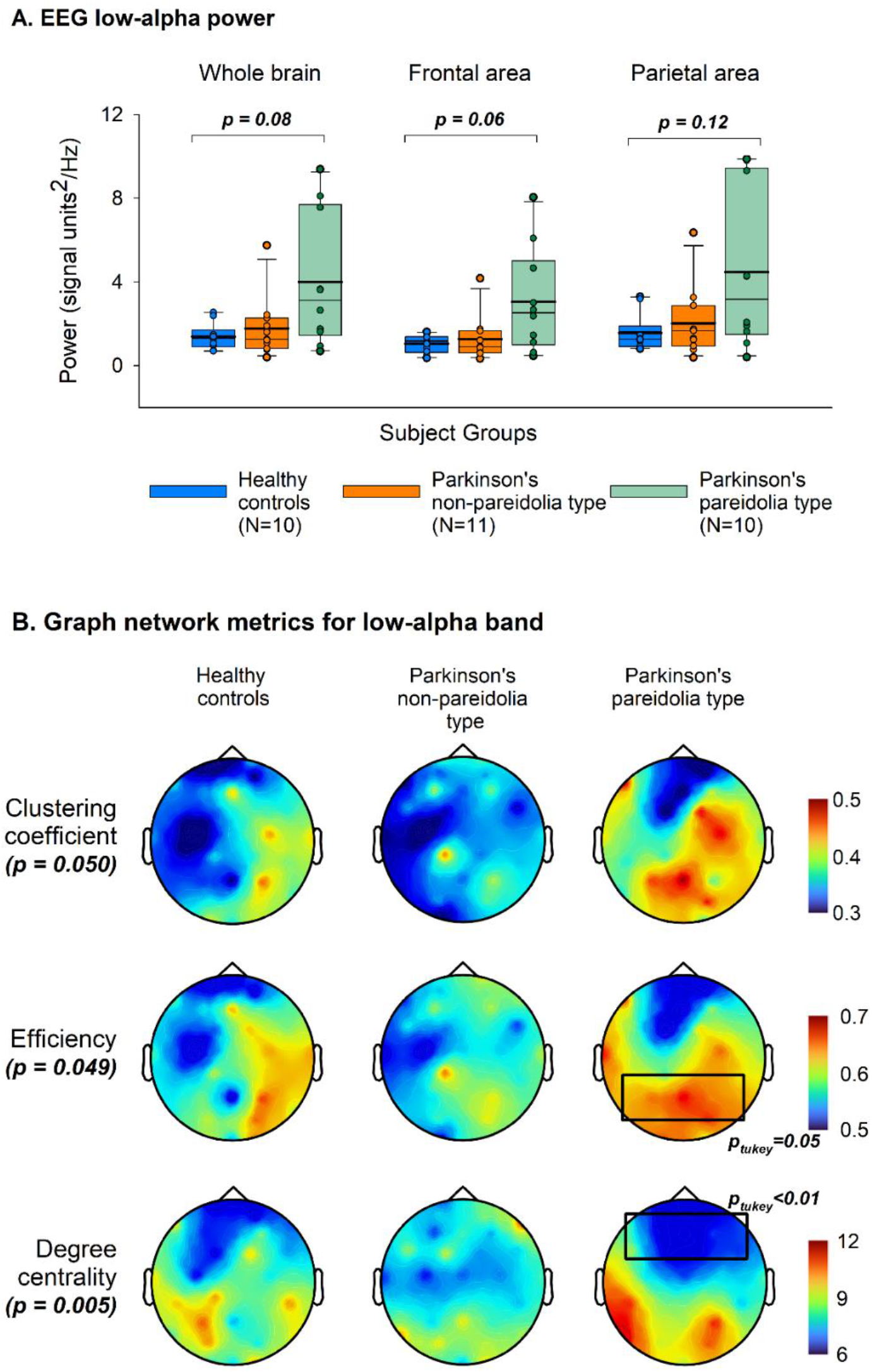
Spectral and topological structures of graph indices for Low-alpha band. (A) Boxplot (with point-plot overlap) shows low-alpha (8 to 10Hz) frequency spectrum for whole brain, frontal and parietal electrodes separately. Within the box, thin black line represents the median whereas thick black line represents the mean. Whole brain refers to full set of 32 electrodes. Frontal electrodes comprised of 13 anterior electrodes, whereas 11 posterior electrodes were included for parietal region. p-values shown are results of ANCOVA analysis (described in Table-2). (B) Topological plot showing significant differences of key graph indices. Warm colors show increased whereas cool colors show decreased nodal values for the respective indices. p-values shown are results of ANCOVA analysis (Table-2). Boxed area (in black) are significant regional post-hoc comparisons.

### Analysis of inter-stimulus intervals

An overview of the evaluation procedure and analysis is shown in Fig.1.

Each NPT image was preceded by a fixation-cross for 2s on the monitor on which the subjects were asked to focus. EEG data were epoched from −2000ms to 0ms, with 0 being the onset of NPT image. To avoid any stimulus-onset related potential from the fixation-cross image, the first 1000ms was discarded. To normalize the recordings, for each subject, the entire epoch was then baseline corrected by its mean value, and wavelet transformed (Morlet) to obtain theta (4-7Hz), low-alpha (8-10Hz) and high-alpha (10-13Hz) frequency bands. In our formulation of graph networks, we analyzed (i) global networks – defined by all 32 electrodes (‘nodes’), and (ii) local measures - pre-defined via 13 frontal and 11 parietal electrodes. Frontal electrodes comprised of Fp1, AF3, F7, F3, FC1, FC5, FC6, FC2, F4, F8, AF4, Fp2 and Fz, whereas parietal area included CP1, CP5, P7, P3, Pz, PO3, PO4, P4, P8, CP6 and CP2 electrodes according to Biosemi cap-coordinate system. This selection was motivated by our pre-defined hypothesis and prior literature(10,34) with respect to the involvement of fronto-parietal networks in target search-and-detect paradigms.

To evaluate graph network properties for each frequency band, we utilized the weighted phase lag index (wPLI) as a measure of network connectivity. True neural interaction between two sources (here, the sensor space) show a coherent relationship when their time-series information is synchronized. wPLI provides a measure of this relationship between the sensor pairs to define cortical signals that are independent of volume conduction and exclude zero-lag connections caused due to spurious uncorrelated noise, thereby improving statistical power to identify phase synchronizations(35,36). Intuitively, wPLI estimates the interaction between electrode pairs and quantifies the functional coupling between brain regions. wPLI values were calculated for theta (4-7Hz), low-alpha (8-10Hz) and high-alpha bands (10-13Hz) using Welch’s power spectrum density(37), and served as ‘edges’ between the network pairs. For each subject, a 32×32 node-to-node adjacency matrix was obtained which were subsequently binarized to create undirected graphs for analysis. A crucial step in binarization is the application of thresholds to the adjacency matrices. Since threshold setting is often arbitrary(38), to obtain consistent results, we calculated the shortest path lengths between interconnected nodes for a range of thresholds and selected the one in which the nodes were connected approx. 99% of the time.

### Graph network indices

Relevant to our hypothesis, we evaluated small-world properties (normalized clustering coefficient and normalized characteristic path length), efficiency (global and local sub-networks) and centrality (degree and betweenness centrality). The clustering coefficient is a measure of functional segregation between locally associated nodes and provides the likelihood that a node is connected by its neighbors. The characteristic path length is the average shortest path length between all possible pairs of nodes within the network and outlines the integration of information from different brain regions. Both the above indices were normalized using 100 random surrogate networks (to obtain Gamma and Lambda values respectively)(39). Efficiency of a graph defines the ability of information exchange within the network. Whereas global efficiency illustrates parallel information processing with a high integration of nodes, local efficiency is the averaged global efficiency within a sub-network of locally selected nodes which illustrates the tolerance of information exchange when a locally connected node is eliminated(27). Centrality measures the effectiveness of a node in information transfer within the network. Here, degree centrality defines the importance of a node(s) and its direct impact on adjacent brain areas. In our formulation of binarized graphs, the degree centrality illustrated the number of connections of each node to all other nodes in the graph. We also evaluated the betweenness centrality which provides a quantification of the node that acts as a bridge between two other connected nodes along its shortest path length. The shortest path length was the smallest number of connections between two nodes and was utilized not only to calculate various network measures, but also to define the sparsity threshold(40).

### Statistical analysis

Differences between demographic and neuropsychological tests were reported using Kruskal Wallis test. For significant p-values, Dunn’s post-hoc tests (for unequal groups) were performed between HC, PDnP and PDP groups. To report differences between patient groups (PDnP and PDP), Student’s t-test or Mann Whitney-U test were performed.

For each spectral and graph network metric, between-group differences were evaluated using ANCOVA analysis on groups (HC, PDnP and PDP) with the addition of age and MoCA scores as nuisance covariates. These covariates were applied based on the fact that cortical activity, especially graph networks, are contingent on age-related cognitive differences(41). Assumptions of normality and homogeneity were performed using Shapiro-Wilk’s test and Levene’s test respectively, with the test statistic set to 0.05. For responses that did not achieve normality, non-parametric ANOVA was performed. Statistical significance was set to p<0.05 with subsequent post-hoc Tukey test performed for each significant dependent variable, adjusted for family-wise comparisons.

Since the aforementioned cortical metrics were calculated specifically for the NPT, we evaluated if these results could be envisioned for other cognitive or visuospatial tests. We therefore implemented a multiple linear regression for the three groups with neuropsychological tests as the clinical dependent variable; and age, EEG spectral power and graph indices as independent variables. Significance for the regression ANOVA model was set to 0.05.

EEG preprocessing and epoching was implemented with an open source toolbox, Brainstorm(42). wPLI indices were calculated using Hermes toolbox(37), the resulting output used as edge weights for network construction using Gretna toolbox(43). Statistical analysis was performed on JASP(44). All other offline analysis and graphing of data were done using Matlab 2017b. Data analyzed in this study will be made available from the corresponding author upon reasonable request.

## Results

Demographic characteristics of participants are summarized in Table-1. In brief, PD patients susceptible to pareidolias (PDP) were older and under-performed on neuropsychological tests.

**Table 1.**
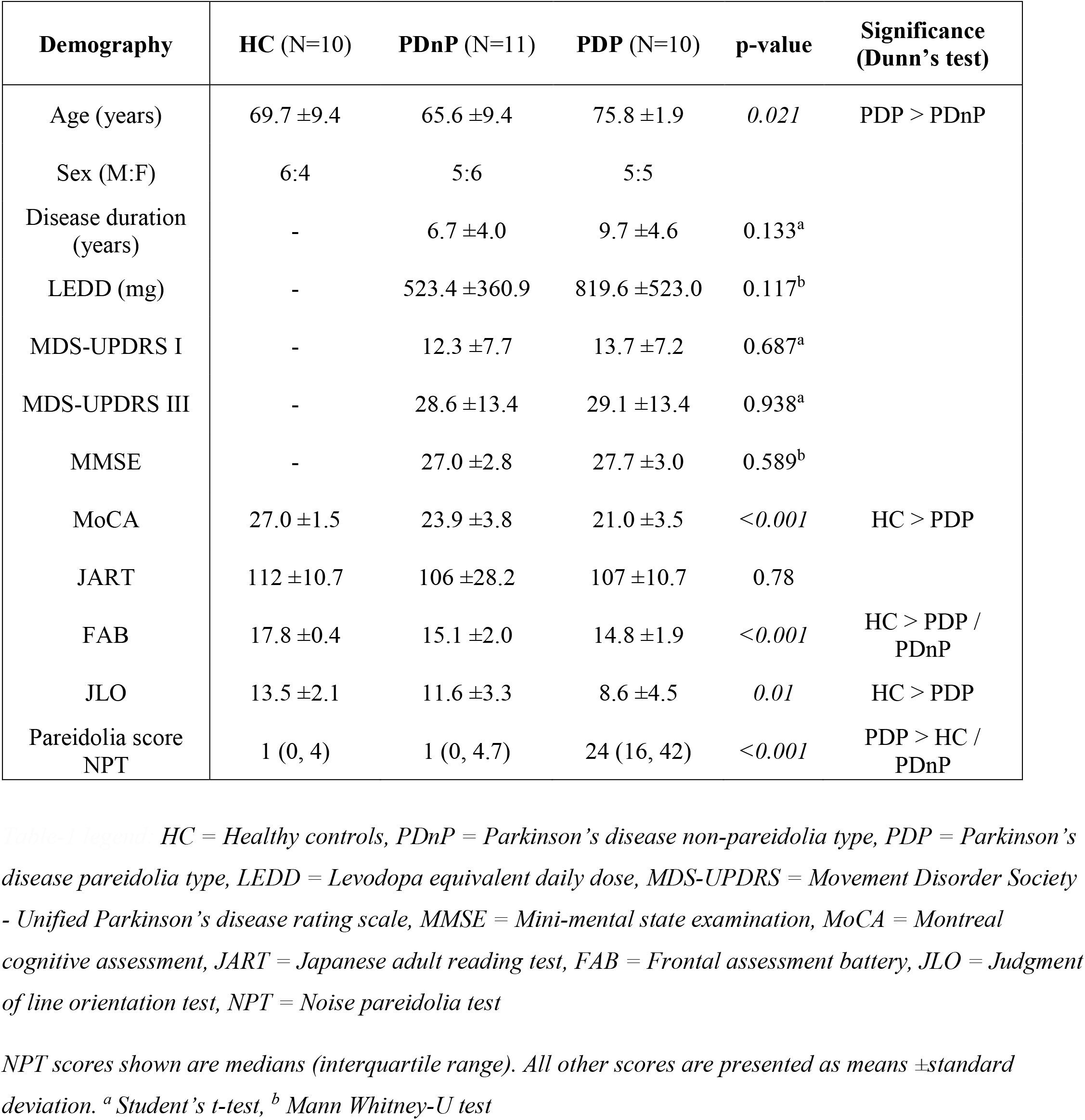
Demographic and neuropsychological assessment.

For EEG analysis of the pre-stimulus phase, a grand total of 780 trials for HC, 840 for PDnP and 755 for PDP group were analyzed. Spectral analysis for theta, low-alpha and high-alpha bands showed no significant differences between the groups. However, low-alpha frequencies in frontal area exhibited a trend towards higher spectral power for the PDP group (p=0.06). Both global and local wPLI showed no significant differences between the groups (Table-2).

**Table 2.** Analysis of covariance on spectral and graph network measures for low-alpha frequency band.

| Measure | Global | Local |  | Post-hoc comparisons |
| --- | --- | --- | --- | --- |
|  |  | Frontal | Parietal |  |
| Mean power (dB)<sup>†</sup> | $F_{(2,28)} = 5.05$<br>$p = 0.08^{\ddagger}$ | $F_{(2,26)} = 2.99$<br>$p = 0.06$ | $F_{(2,28)} = 2.27$<br>$p = 0.12$ | - |
| Mean wPLI | $F_{(2,26)} = 0.25$<br>$p = 0.77$ | $F_{(2,26)} = 0.63$<br>$p = 0.54$ | $F_{(2,26)} = 0.10$<br>$p = 0.90$ | - |
| Normalized clustering coefficient | **$F_{(2,26)} = 3.38$**<br>**$p = 0.050^*$** | $F_{(2,26)} = 0.06$<br>$p = 0.94$ | $F_{(2,26)} = 3.14$<br>$p = 0.06$ | PDP > PDnP,<br>$p_{\text{tukey}} = 0.06$ |
| Normalized path length | $F_{(2,26)} = 1.52$<br>$p = 0.24$ | $F_{(2,26)} = 0.83$<br>$p = 0.45$ | $F_{(2,26)} = 0.44$<br>$p = 0.65$ | - |
| Efficiency | $F_{(2,26)} = 0.16$<br>$p = 0.85$ | $F_{(2,26)} = 0.98$<br>$p = 0.34$ | **$F_{(2,26)} = 3.39$**<br>**$p = 0.049^*$** | PDP > PDnP,<br>$p_{\text{tukey}} = 0.050$ |
| Degree centrality | $F_{(2,26)} = 0.12$<br>$p = 0.89$ | **$F_{(2,26)} = 6.47$**<br>**$p = 0.005^*$** | $F_{(2,26)} = 1.63$<br>$p = 0.21$ | PDP < PDnP,<br>$p_{\text{tukey}} = 0.005$ |
| Betweenness centrality | $F_{(2,26)} = 0.24$<br>$p = 0.78$ | $F_{(2,26)} = 0.97$<br>$p = 0.39$ | $F_{(2,26)} = 0.35$<br>$p = 0.70$ | - |

<sup>†</sup>Addition of covariates (Age and MoCA) failed tests for equal variances. One-way ANOVA (without covariates) showed significant differences in mean power at global level ( $p = 0.037$ ) and for frontal electrodes ( $p = 0.020$ ), with post-hoc tests showing mean power for PDP group $>$ HC or PDnP groups.
<sup>‡</sup>Non-parametric Kruskal-Wallis ANOVA
\*For all significant comparisons, post-hoc tests (Tukey) are shown in the right-most column.

Calculating the shortest path lengths for a series of ten sparsity thresholds ranging from 0.05 to 0.5, incrementing every 0.05, we fixed the threshold at 0.3 wherein the nodes were connected approx. 99% of the time (Fig.1, Sparsity threshold). Subsequent ANCOVA analysis of the graph networks revealed significant differences for low-alpha frequency band (Table-2), with no differences in theta and high-alpha activity (Supplementary Table-1). In general, low-alpha band changes between HC and PDnP groups were inconsequential. However, PDP group showed a statistically higher normalized clustering coefficient (PDP_Gamma_= 1.04±0.03 vs. PDnP_Gamma_ =0.94±0.03 or HC_Gamma_ =0.93±0.03, adjusted means ±error), with no significant differences in the normalized path lengths. Locally, parietal electrodes showed a higher nodal efficiency which served to demonstrate the higher information exchange of the interconnected network in the parietal area during the pre-stimulus phase. Furthermore, PDP group showed significantly low degree centrality in frontal electrodes suggestive of a dispersed or disconnected network. Topological illustration of graph indices is shown in Fig.2.

Multiple linear regression between clinical variables and spectral/network metrics are shown in Table-3. Results of regression analysis indicated that the spectral and graph indices were significantly correlated to pareidolia scores (R^2^=0.43, *p=0*.*011*) and weakly to FAB scores (R^2^=0.20, p=0.052). The regression coefficients for the independent variables are described in Supplementary Table-2.

**Table 3.**
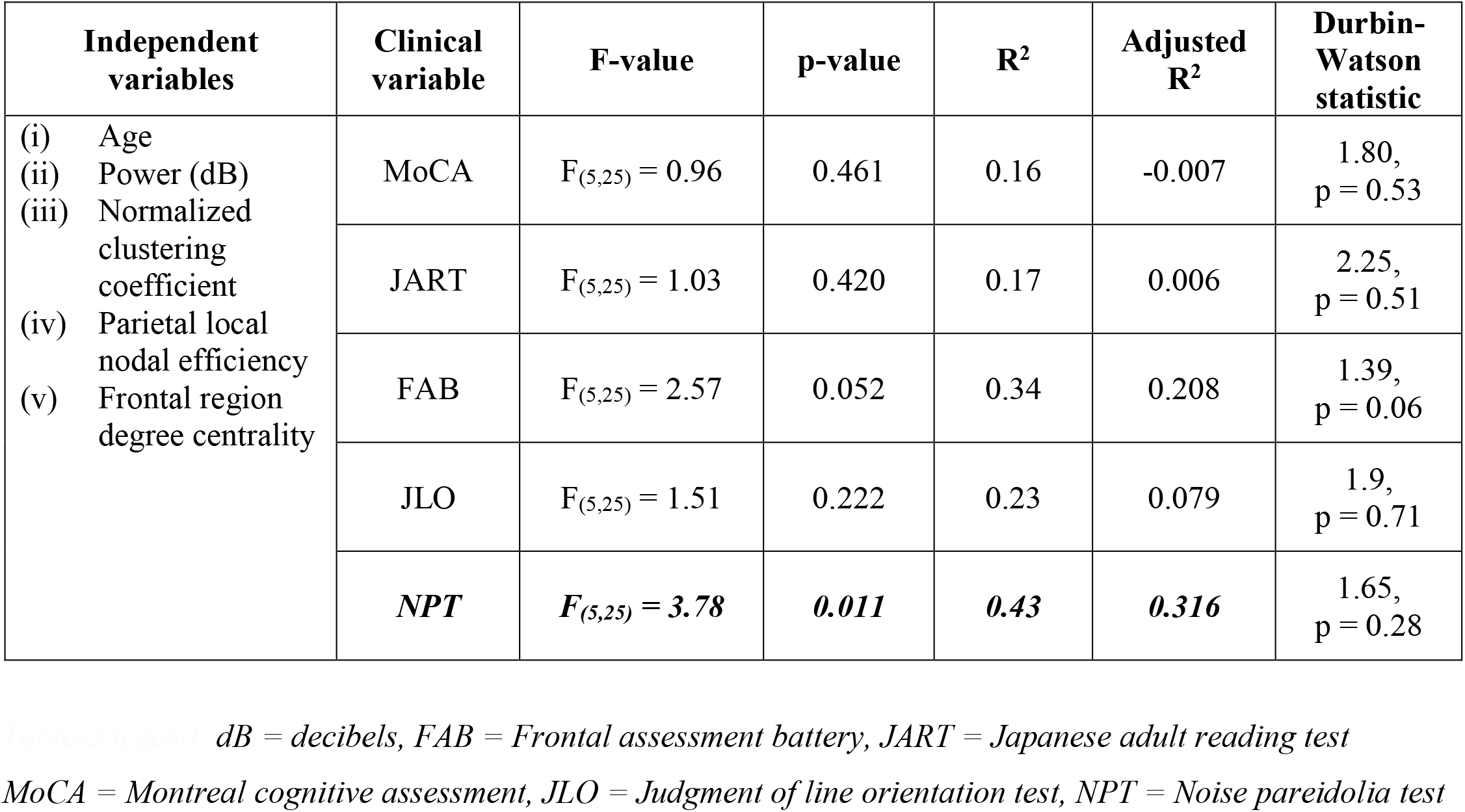
Multiple linear regression of low-alpha spectral and graph indices for neuropsychological tests.

## Discussion

We investigated how large-scale network organization is altered prior to stimulus onset in PD patients susceptible to pareidolias. In patients who demonstrate pareidolic misperceptions, we found that: (i) the pre-stimulus low-alpha power tends to be higher in frontal cortex, (ii) there are notable changes in global and local network properties during anticipation of stimuli, and (iii) the spectral and network indices are characteristic of pareidolic expression on the noise pareidolia test (NPT).

Face detection on the NPT necessitates an active search-and-detect effort requiring configural face processing and a top-down integration of facial features(10). While these images serve as sensory deprived ambiguous stimuli, a subset of PD patients consciously misperceive noisy patches as faces, likely due to abnormal perceptual biases(13). In our experiment during the pre-stimulus phase, this misrepresentation was observed as a near-significant increase in low-alpha spectral power in the frontal electrodes. In literature, alpha power codes stimulus identity(45). Conscious perception of low-level information reflecting top-down visual processing has been attributed to higher pre-stimulus low-alpha power(46). Anticipation of relevant stimuli increases alpha activity, its strength proportional to the saliency of the stimuli(47). Given that cortical low-alpha power represents the attentional dynamics within the cortico-cortical loops(45,48), our findings are suggestive of a diversion of attentional resources towards internal processing to code non-relevant stimulus identity in patients susceptible to pareidolias. However pre-stimulus low-alpha power is increased globally i.e. in frontal, parietal and occipital electrodes when targets are actively perceived(46). Our results partially support this finding since we did not find any differences in the posterior parietal electrodes which have a critical role in pre-stimulus internally generated processes(49). This discrepancy could be attributed to a diverse pathology seen in pareidolias, which are known to occur at different stages of PD involving both cortical (frontal or parieto-occipital cortex) and subcortical structures (upper brainstem or thalamus)(7).

Pareidolic misperceptions, like hallucinations, are an eventuality of global cortical network dysfunction(8). Pertaining to global network level deficits in PD, we explored small-world properties which are commonly affected in neuropsychiatric diseases(39). The increased normalized clustering coefficient reported in the PDP group suggests a heightened integration of information during pre-stimulus phase. Visualization of the topology showed a delineation of clusters in the parietal cortex (Fig.2). Furthermore, these posterior nodes also revealed a high nodal efficiency when compared with HC or PDnP groups. As a proxy to small-worldness(50), these indices demonstrate an efficient information transfer within the parietal network. Within the framework of low-alpha band frequency, our findings suggests a reduced disengagement of posterior cortex during non-relevant stimulus anticipation(15). This cortical signature in the posterior electrodes reflecting focused intentional awareness(49) is a persisting feature for ambiguous stimuli, since similar findings have also been observed in PD patients with hallucinations via bi-stable percept paradigms that demonstrate the altered communication between the default mode network (DMN) and the dorsal attentional network (DAN)(28,51).

The degree centrality is an index of how well the nodes are connected, whereas betweenness centrality is a measure of the node connected to other independent node clusters(40). We observed that patients experiencing pareidolias had significantly lower degree centrality in the frontal electrodes without any change in betweenness centrality. Intuitively, this signifies a higher number of discrete nodes with low connections within the frontal cortex. According to scale-free distributions, in sparsely connected networks, these nodes represent a low-linkage with an absence of hub characteristics(52). Psychophysiologically, with a concomitant high frontal spectral power, this could imply the maintenance of cortical excitability within the network to process imminent visual stimuli irrespective of its saliency(46). The fallout of this process is a breakdown of communication between the DMN and DAN that modulates visuo-perceptual priors(28), disordered frontal attention networks that create an abnormal competition with weak stimuli(53) or an increased dependence on top-down signals leading to the development of misperceptions(54).

We found that the spectral and graph indices were significantly correlated only to pareidolia scores in our participant groups. Our selection of neuropsychological tests was to examine the effect of altered networks in visuo-perception that overlap with cognitive (MoCA, JART), executive (FAB), visuo-spatial and visuo-construction function (JLO). While FAB showed a trend towards significance, the less than significant findings from JLO were notable since posterior parietal cortex lesions often show lower scores on JLO(55). It is therefore likely that the high clustering coefficient and nodal efficiency we observed in the parietal electrodes could be an epiphenomenon observed due to stimulus anticipation process. Since regression did not affect the outcome of other neuropsychological tests, the above findings signify a higher influence of fronto-cortical network structure on the outcome of pareidolic misperception. Regardless, profiling disease pathology in neuropsychiatric disorders in terms of increased or decreased connectivity purely on the basis of graph theory must be treated with caution due to overlaps in domain-specific tasks with different brain areas(50).

### Limitations

There are some limitations to this study. The scope of this report was mainly to demonstrate the cortical dynamics via EEG though other behavioral measures may be relevant. Eye-tracking metrics such as change in low-frequency pupillary power is suggested to be crucial during stimulus anticipation in patients experiencing pareidolias(56). The effect of negative mood is also known to affect the outcomes of the NPT(13). We also avoided sub-classifying the pre-stimulus responses into face, noise, pareidolias and missed images due to the skewed nature of the NPT (fewer face stimuli than noise). Furthermore, the saliency of Mooney face stimuli within the NPT was not trivial, and considering the length of the experiment, some degree of temporal variability of the EEG was unavoidable(57). The effect of levodopa(58) was not systematically studied except that the NPT was always performed during ‘ON’ state of medication. We also did not report here crucial subjective experiences of patients (images that were responded to as animals etc.) which may be relevant to the concept of illusionary responses. Therefore, more data by increasing sample sizes are necessary to understand conclusively the effects of pareidolia.

Statistically, the data distribution followed gaussian allowing us to control our analysis methodologies for age and cognitive levels. Although addition of other covariates (eg. JLO, FAB) increased the statistical significance of the tests (data not shown), we avoided their inclusion due to a paucity of prior literature on their effects on graph networks. Our mode of comparing normal and diseased networks of similar size by fixing thresholds for binarized graphs is in line with prior literature although weighted graphs could be an informative substitute to compare such conditions. Addressing these issues in the future would be beneficial not only for cross-sectional studies but also for studies that focus on the longitudinal evolution of pareidolia in the culmination of hallucinations/psychosis in PD.

## Conclusions

Pareidolias in PD are a consequence of an abnormal top-down modulation of visual processing(10) which are defined by frontal low-alpha spectral and network alterations in the pre-stimulus phase due to a dissonance between patients’ internally generated mental-processing with external stimuli. As a surrogate of visual hallucinations, our result highlights the impact of pareidolias on the frontal cortex in susceptible PD populations. Further work is needed to ascertain the burden of cortico-subcortical structures in the pathogenesis and maintenance of such symptoms in PD.

## Supporting information

STROBE_Reporting guidelines

## Data Availability

Data analyzed in this study will be made available from the corresponding author upon reasonable request.

## Acknowledgements

We express our sincere gratitude to the patients who participated in this study, and to Mr. Toshiaki Fujimoto and the members of the Minoh Senri-Chuo Rotary Club, Japan for their undivided support. We also thank Dr. Maki Suzuki and Dr. Hiroyuki Watanabe for their advice during the development process.

## Authors’ Roles

Conceptualization - G.S.R., Y.K., T.S., N.H., E.M. and H.M.

Methodology - G.S.R., T.N. and Y.K.

Software, Validation - G.S.R., Y.K. and T.S.

Formal analysis, Data curation – G.S.R., N.H., T.N. and Y.K.

Writing – original draft preparation - G.S.R.

Writing – review & editing – Y.K., T.S., T.N., N.H., M.M., E.M and H.M.

Supervision, Funding acquisition – G.S.R. and H.M.

