## Supplemental files for "Pre-stimulus low-alpha frontal networks characterize pareidolias in Parkinson’s disease"

**Supplementary data**

**Supplementary methodology:**

For our study, Mooney faces used had a shadow-effect that were slightly more ambiguous (Supplementary Fig. 1) than on the paper-based test. We performed this step to (i) avoid the floor-effect with low pareidolia scores on the NPT seen in PD patients(1,2), (ii) increase the ambiguity of the images without altering its visuo-spatial characteristics of the face stimuli(3). Mooney faces are low-information, two-tone pictures of faces to test face perception – a sorting task in adults which requires configural face processing and is dependent on knowledge-based integration of facial features(4,5). 2D Mooney face dataset were obtained from the Psychological Image Collection at Stirling webpage made available for public use at pics.stir.ac.uk. For statistical analysis, the projected sample size needed for a three group ANOVA with minimum detectable mean level = 0.3, expected standard deviation of residual = 0.15, alpha = 0.05 and power = 0.95 was N (participants) = 9.

**Supplementary Figure 1:** Example image of a noise pareidolia test with (A) Mooney face embedded, and (B) pure noise.

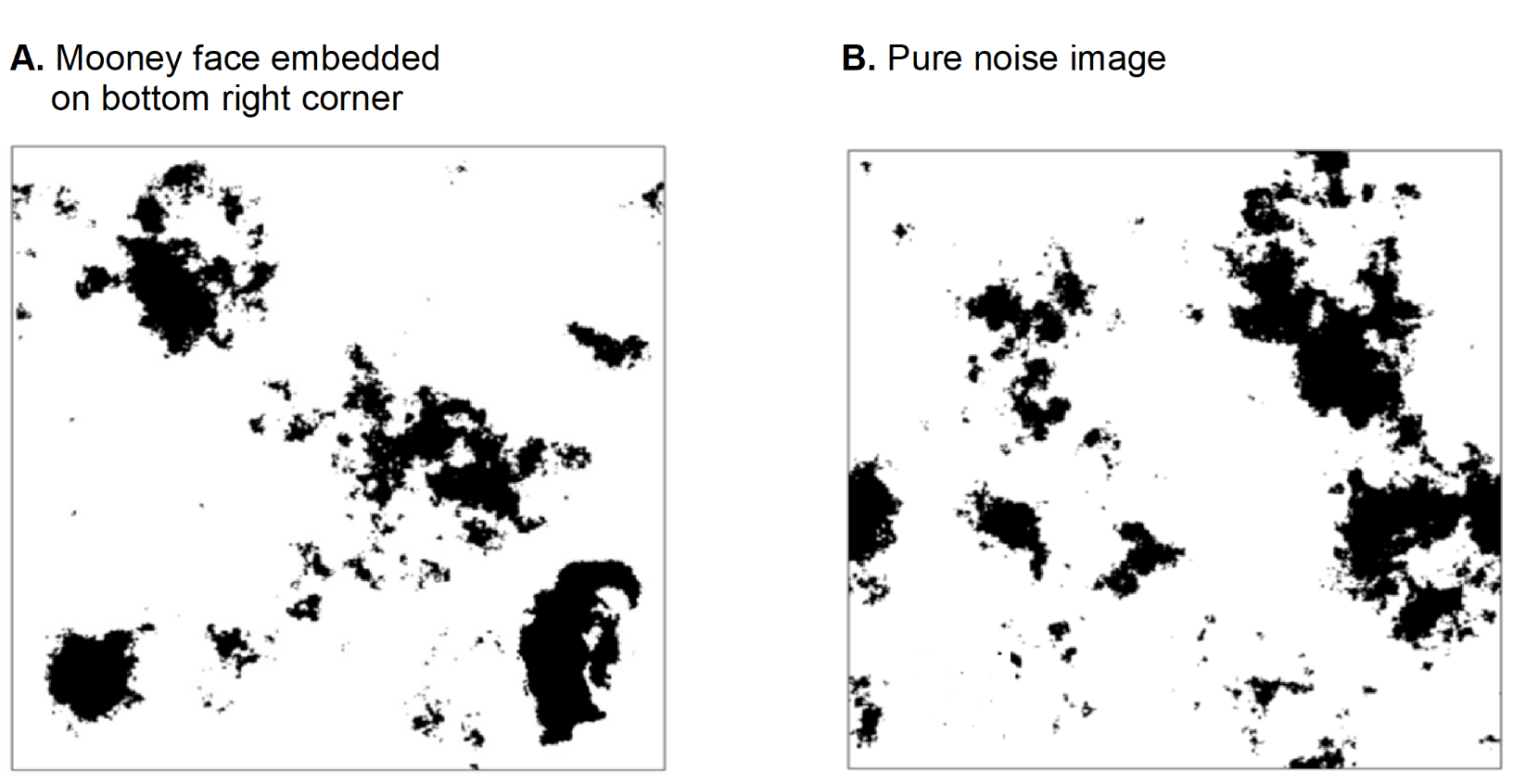

**Supplementary Table-1:** Spectral and graph network measures for theta and high-alpha frequency bands

| **Frequency** | **Graph index** | **Global** | **Local** | |
| --- | --- | --- | --- | --- |
|  |  |  | ***Frontal*** | ***Parietal*** |
| Theta  (4-7 Hz) | Mean power (dB) | F_(2,26)_ = 1.39  p = 0.26 | F_(2,26)_ = 1.48  p = 0.24 | F_(2,26)_ = 0.61  p = 0.55 |
|  | Mean wPLI | F_(2,26)_ = 2.58  p = 0.09 | F_(2,26)_ = 0.27  p = 0.76 | F_(2,26)_ = 3.62  p = 0.051 |
|  | Normalized Clustering coefficient | F_(2,26)_ = 0.23  p = 0.79 | F_(2,26)_ = 0.86  p = 0.43 | F_(2,26)_ = 0.51  p = 0.60 |
|  | Lambda (Normalized path length) | F_(2,26)_ = 0.09  p = 0.91 | F_(2,19)_ = 0.31  p = 0.73 | F_(2,25)_ = 0.76  p = 0.48 |
|  | Efficiency | F_(2,26)_ = 0.95  p = 0.40 | F_(2,26)_ = 0.34  p = 0.71 | F_(2,26)_ = 0.24  p = 0.78 |
|  | Degree centrality | F_(2,26)_ = 0.40  p = 0.67 | F_(2,26)_ = 0.92  p = 0.41 | F_(2,26)_ = 0.05  p = 0.95 |
|  | Betweenness centrality | F_(2,26)_ = 0.23  p = 0.79 | F_(2,26)_ = 1.75  p = 0.19 | F_(2,26)_ = 1.09  p = 0.35 |
| High Alpha  (10-13 Hz) | Mean power (dB) | F_(2,26)_ = 1.62  p = 0.21 | F_(2,26)_ = 2.09  p = 0.14 | F_(2,26)_ = 0.34  p = 0.71 |
|  | Mean wPLI | F_(2,26)_ = 0.58  p = 0.57 | F_(2,26)_ = 0.87  p = 0.43 | F_(2,26)_ = 1.20  p = 0.31 |
|  | Normalized Clustering coefficient | F_(2,26)_ = 0.20  p = 0.82 | F_(2,26)_ = 0.34  p = 0.71 | F_(2,26)_ = 0.53  p = 0.59 |
|  | Normalized path length | F_(2,26)_ = 0.21  p = 0.81 | F_(2,21)_ = 2.60  p = 0.09 | F_(2,26)_ = 5.15  p = 0.013 |
|  | Efficiency | F_(2,26)_ = 0.41  p = 0.67 | F_(2,26)_ = 0.07  p = 0.93 | F_(2,26)_ = 0.15  p = 0.86 |
|  | Degree centrality | F_(2,26)_ = 1.10  p = 0.35 | F_(2,26)_ = 1.54  p = 0.23 | F_(2,26)_ = 2.86  p = 0.075 |
|  | Betweenness centrality | F_(2,26)_ = 1.8  p = 0.18 | F_(2,26)_ = 0.08  p = 0.92 | F_(2,26)_ = 1.02  p = 0.38 |

*Legend:
dB = power spectrum in decibels
wPLI = weighted phase lag index*

**Supplementary Table-2:** Regression coefficients for the noise pareidolia test for low-alpha band frequency (8 to 10 Hz)

| **Independent Variables** | **Unstandardized** | **Standard error** | **Standardized** | **t-value** | **p-value** |
| --- | --- | --- | --- | --- | --- |
| Age | 0.44 | 0.29 | 0.24 | 1.51 | 0.142 |
| Mean power | 1.45 | 0.73 | 0.32 | 1.97 | 0.059 |
| Normalized clustering coefficient | 61.23 | 26.93 | 0.35 | 2.27 | ***0.032*** |
| Parietal nodal efficiency | 15.57 | 39.94 | 0.07 | 0.39 | 0.701 |
| Frontal degree centrality | -1.52 | 2.32 | -0.13 | -0.65 | 0.518 |
